# The Veterans ECS21 (Exploring Cannabigerol for Sleep) Study: A protocol for the first randomized clinical trial of Cannabigerol (CBG), a decentralized study designed for U.S. Veterans in California

**DOI:** 10.1101/2023.03.13.23287212

**Authors:** Courtney E Webster, Chris R Emerson, EJ Daza

**Author notes:** **Corresponding author** Courtney Webster, nymbly, Seattle, WA, USA. **Author ORCIDs and emails.** Chris R Emerson Courtney E Webster EJ Daza.

## Abstract

The Veterans ECS21 (Exploring Cannabigerol for Sleep) Study is the first clinical trial of Cannabigerol (CBG), a non-psychoactive cannabinoid (ClinicalTrials.gov Identifier: NCT05088018). This study investigates if CBG improves sleep for U.S. Veterans in California using a fully remote (decentralized), randomized, placebo-controlled, triple-blind, pre–post study design. Over 100 participants will be recruited through Veterans advocacy organizations, email campaigns, and point-of-sale displays at cannabis dispensaries. Participants will be provided with a study application to assess sleep quality, quality of life, and PTSD-related symptoms. A Fitbit Inspire 2 will be used to collect sleep, activity, and cardiovascular biometrics. After a two-week run-in phase, participants will begin a four-week treatment phase with a dose escalation from 25 mg daily CBG (CBG Protab™ by LEVEL) up to 50 mg CBG daily. The present paper describes how we designed this study to overcome specific challenges. We held participant insight panels to learn Veterans’ lived experience and design ways to lower their obstacles to clinical research participation. We designed digital engagement strategies to maintain high participation and retention for a fully remote (decentralized) study, selected fit-for-purpose passive data collection mechanisms, and addressed common reproducibility issues in cannabis research.

## Introduction

While one out of three people report trouble sleeping in their lifetime, the Veteran population is up to six times more impacted by sleep-related issues than the general population.^1^ The United States Veterans Health Administration (VHA) considers sleep issues among Veterans a healthcare crisis. From 2012 to 2018, insomnia diagnoses for Veterans nearly doubled, and sleep-related breathing disorders increased four-fold.^2^ Veterans diagnosed with sleep disorders commonly have comorbidities including obesity, diabetes, congestive heart failure, depression, post-traumatic stress disorder (PTSD) or traumatic brain injury.^2^

Hemp and cannabis are most commonly used to manage pain, anxiety, and sleep problems.^3,4^ Delta-9-tetrahydrocannabinol (_Δ_9-THC) is the primary psychoactive component of cannabis,^3^ and makes up almost 95% of cannabis sales (**Supplementary Table 1**),^5^ while less psychoactive cannabinoids such as cannabinol (CBN) and non-psychoactive cannabinoids such as cannabidiol (CBD) and cannabigerol (CBG) are gaining in popularity for both recreational and clinical use. Growing research indicates that cannabis is used by Veterans with PTSD to cope with sleep disturbances and other PTSD symptoms.^6,7^ Whole plant cannabis and varying ratios of _Δ_9-THC/CBD have been studied for their effects on sleep.^4,8-12^ While anecdotally, CBG users report improved sleep, no clinical studies exist to assess its efficacy. CBG exhibits similar activity and affinity characteristics as _Δ_9-THC and CBD on cannabinoid receptors, but has unique high (nanomolar to sub-nanomolar) affinity as an _α_-2 adrenoceptor agonist.^13^ Other _α_-2 agonists such as FDA-approved clonidine exhibit antihypertensive, sedative, and analgesic activity.^14^ Clonidine has demonstrated clinical benefit for insomnia in children with attention deficit hyperactivity disorder (ADHD) as well as some efficacy in improving sleep in adults with PTSD.^15^

## Objective

The goals for this study are to assess how a daily dose of orally self-administered CBG impacts sleep and quality of life for California Veterans. This pilot study is crucial to understanding the potential risks and benefits of CBG as a treatment for sleep issues. It will also establish feasibility, precedence, and generate preliminary findings that can be used for future trials.

The primary objective of this study is to estimate the mean effect of CBG on the pre–post change (i.e., change from the pre-treatment run-in period to four weeks after the start of treatment) on sleep using the Sleep Problems Index II subscale of the Medical Outcomes Study Sleep Scale (MOS-SS SPI-II).^16-18^ We chose the MOS-SS SPI-II as it was validated using a large, heterogeneous population of adults (n=3,445) with chronic conditions that exhibited high incidence rates of insomnia and sleep disturbances,^16,18^ and is now a well-established instrument in assessing sleep quality in the U.S. veteran populations.^19-22^ While initially both the MOS-SS and the MOS-SS SPI-II subscale evaluated self-reported sleep quality over the past 4 weeks (i.e., using a past 4-week recall interval),^16-18^ modifications to evaluate sleep across shorter periods (e.g., over a 1 or 2-week time frame) are feasible and validated.^19,23^ For the present study, we chose a 2-week timeframe.

For our secondary endpoint, we wanted to prioritize a measurement that mattered to our participants – or, in the Digital Medicine (DiMe) Society’s terminology, a “meaningful aspect of health”.^24^ We felt a quality of life (QoL) assessment would allow us to evaluate the holistic effect sleep has on daily functioning, well-being, emotional and community engagement, and other parameters. While many QoL questionnaires focus only on physical function, the World Health Organization Disability Assessment Schedule, version 2.0 (WHODAS-2.0, https://www.who.int/) is applicable across all health states, including drug and alcohol problems, various mental health conditions (PTSD, depression, anxiety, and schizophrenia) as well as physical conditions.^25-28^ The conventional WHODAS-2.0 questionnaire contains 36 items assessing disability and limitations experienced over the preceding 30 days across six domains: cognition, mobility, self-care, getting along with others, life activities, and participation (six items for each domain) and has been used in previous studies in US Veteran populations.^29,30^ An abbreviated 12-item self-report version (WHODAS-2.0–12) in which each domain has two items is also available and commonly used.^25,28^ For simplicity we also chose this 12-item version.

One exploratory objective is to evaluate the effective dose by comparing the mean effect of the initial dosing regimen (i.e., after the first two weeks of the treatment period) to that of the higher dosing regimen (i.e., at the end of all four weeks of the treatment period). Other exploratory objectives are to evaluate how CBG impacts sleep, activity, and heart rate biometrics as measured by a Fitbit Inspire 2. It is interesting to note that even a modest increase in physical activity can decrease depression in adults.^31^ Therefore, collecting activity metrics could provide some data to be used for exploratory analysis related to activity, sleep, and mental health. Lastly, as other _α_-2 agonists exhibit hypertensive activity, we could evaluate if CBG and or CBG dose effects heart rate. The additional activity metrics and heart rate could be collected with no added subject burden.

Lastly, we will monitor changes in PTSD symptoms, given the high precedence of PTSD co-occurring (if not directly responsible for) sleep-related issues in Veterans. PTSD symptoms will be evaluated using the PTSD Checklist for DSM-5 (PCL-5).^32,33^

The PCL-5 is a brief, self-report symptom checklist that assesses the 20 symptoms of PTSD outlined in the Diagnostic and Statistical Manual-5 (DSM-5) and was designed to assess symptom changes during and after treatment in addition to screening.^32,33^

Our pre–post study design allows us to measure the participant’s experience with both patient-reported outcomes (PROs) before and after taking the CBG tablet intervention. We will also study a placebo effect by comparing changes in subjective reporting between the intervention and placebo arms of the study.

## Study design challenges

We identified number of study design challenges to overcome in order to make the mission and motivations of this study a reality.

### Challenge #1: Recruiting and engaging a skeptical Veteran community

First and most importantly, this research was designed to support the Veteran community. C.R. Emerson, the founder of Metta Medical and the study sponsor, is a Veteran. We wanted to design recruitment techniques that would reflect the lived experiences of our study population. The Veteran community is skeptical about research, and many Veterans report negative experiences when receiving medical care. The fear of getting “5150’d” (e.g., placed in an involuntary psychiatric hold)^34^ keeps many Veterans from answering questions about sensitive topics honestly.^35^ We planned to ask about study participant’s cannabis use, drug and alcohol use, and mental health medications and knew we needed to demonstrate our commitment to their privacy in order to establish their trust.

### Challenge #2: Recruiting and retaining a representative study population

The VHA has implemented numerous innovations to improve the quality and accessibility of sleep care services for Veterans (https://www.veteranshealthlibrary.va.gov/).^2,36,37^ And yet, access to diagnostic sleep testing, sleep specialists, and treatment is limited. This access was already difficult for rural Veterans and exacerbated during the COVID-19 pandemic. We desired a decentralized study design to allow equitable access – however, a remote study design presents its own challenges with retention. A recent cross-study evaluation of 100,000 participants in eight remote digital health studies reported a median participant retention of only□5.5 days.^38^ We needed to design techniques that would recruit and engage participants for at least six weeks.

### Challenge #3: Minimizing or avoiding common confounders in cannabis research

Reproducibility in cannabis studies is challenged by the pharmacological diversity of cannabinoids when extracted from the plant, as different strains have variable concentrations of active constituents. Therapeutic efficacy also depends on a person’s current tolerance of cannabis,^39^ and varies due to the method of administration (inhalable, oral, topical, sublingual, rectal/vaginal). These and other issues make it challenging for researchers to reproduce results. As this is the first reported clinical study of CBG, we wanted to avoid these challenges where possible so the field can continue to build on these results.

### Challenge #4: Effectively measuring a change in sleep quality: Endpoint & Digital Health Technology (DHT) design

Sleep is challenging to study, as it is easily impacted by physiological and environmental circumstances. We planned to include PRO questionnaires, but also wanted to include a digital clinical measure of sleep. Polysomnography (PSG) was out of scope for this study – it was simply not feasible to require proximity to a sleep lab as one of our inclusion criteria – so we desired remote monitoring technology that participants could use at home. We used DiMe’s “V3” Framework,^40^ and the EVIDENCE checklist^41^ to guide our fit-for-purpose device selection process.

## Methods

### Participant Panels

We recruited Veterans to join a Participant Panel through our co-sponsor, the Veterans Cannabis Group. We did not have specific inclusion/exclusion criteria to join the participant panels, except for having Veteran status and a willingness to volunteer their time. We had five attendees distributed across three virtual sessions. Attendees included two female and three male participants; most of the attendees were in the 30s–40s age range.

Each panel was moderated by C.E. Webster using a prepared moderator guide. The panel began with an introduction and the goals of the panel. We assured them of their privacy and asked their permission to record the session. We asked participants to tell us about themselves and to describe a typical day in their lives.

We then showed them a study recruitment advertisement, a few questions from the PCL-5 to gauge their comfort with answering sensitive questions on a mobile device (**Supplementary Table 2: Participant Panel Excerpt**), and our proposed daily survey Self Assessment Manikin (SAM) and requested feedback.

### Inclusion and Exclusion criteria

A full list of inclusion and exclusion criteria are shown in **Table 1**. It is important to have our participant population be as representative as possible to the generalized Veteran population. We discuss here how we considered three indications that have higher prevalence in Veteran than non-Veteran populations and what effect, if any, these indications should have on our study population.

First, we considered obstructive sleep apnea (OSA). Our screening questionnaire will ask prospective participants if they have been observed gasping, choking, or having stopped breathing in their sleep and if they have been diagnosed with sleep apnea. Participants who respond ‘yes’ will be excluded if they have not been using a continuous positive airway pressure (CPAP) device for at least four weeks prior to starting the study. If a participant reports using a CPAP device for at least four weeks, we infer that the obstructive element of OSA could be mitigated.

Given the high prevalence of PTSD co-occurring (if not directly responsible for) sleep-related issues in Veterans,^42^ it would be impractical to exclude Veterans on the basis of a PTSD diagnosis. Instead, we include a PTSD-specific questionnaire (the PCL-5) to explore any differences in PTSD symptoms over the course of the study.

Lastly, it would not be feasible or representative to restrict pharmacological use, including mental health-related or even sleep-related medication. We will exclude participants that have stopped, started, or changed medications two months prior to beginning the study. We ask participants to confirm to the best of their ability that current medications would be continued for the duration of the study. We will query participants on their daily use of certain medications as well as whether they’ve stopped, started, or changed the dosage of prescribed medications during the study to account for any mediators or moderators of CBG’s effects

Finally, following initial eligibility screening, we included a MOS-SS SPI-II score threshold of _≥_30 as a proxy indicator of sleep disturbance in order to ensure that we are starting with a population who were experiencing some degree of sleep-related issues prior to the beginning of the study. This threshold is broadly comparable to that used in previous studies in US veterans (where a threshold of _≥_35 was used).^19,20^

### Recruitment

Participants will be recruited across California through Veteran’s associations and advocacy groups, such as the Veterans Cannabis Group, Tactical Patients Group, Operation EVAC, the Santa Cruz Veterans Alliance, and others. Potential participants will be made aware of the study via a number of routes including electronic communication via email, printed materials, and study advertisements at point-of-sale dispensary locations.

### Randomization and Sample Size

We will assign each of the 100 (or more) enrolled participants equally across both treatment arms via stratified block randomization. Two randomization lists will ensure that treatment assignment is balanced according to reported sex to be as consistent as possible with our target population. The female-to-male ratio in the general population of Veterans is 1:9.

We will enroll at least 50 participants per arm and allow for at most 40% attrition down to 30 participants per arm by the end of the study. This provides at least 80% power to discern a true mean difference between the two groups, according to the Analysis Plan described herein.

### Study design

This study is designed to be entirely remote (decentralized), from eligibility screening and consent through to the completion of study tasks. With this design, participants can successfully enroll and participate without visiting a clinical research site.

Following screening and registration, participants will be randomized to either the active treatment or placebo control arm and then enter a two-week run-in period. During the run-in period, participants will begin answering questionnaires assessing their sleep quality, quality of life, and PTSD symptoms. They will answer a daily morning questionnaire about their sleep as well as a nightly questionnaire about their day (**Figure 1: Study Schedule of Assessments** and **Supplementary Table 3: Study Instruments**).

**Figure 1.**
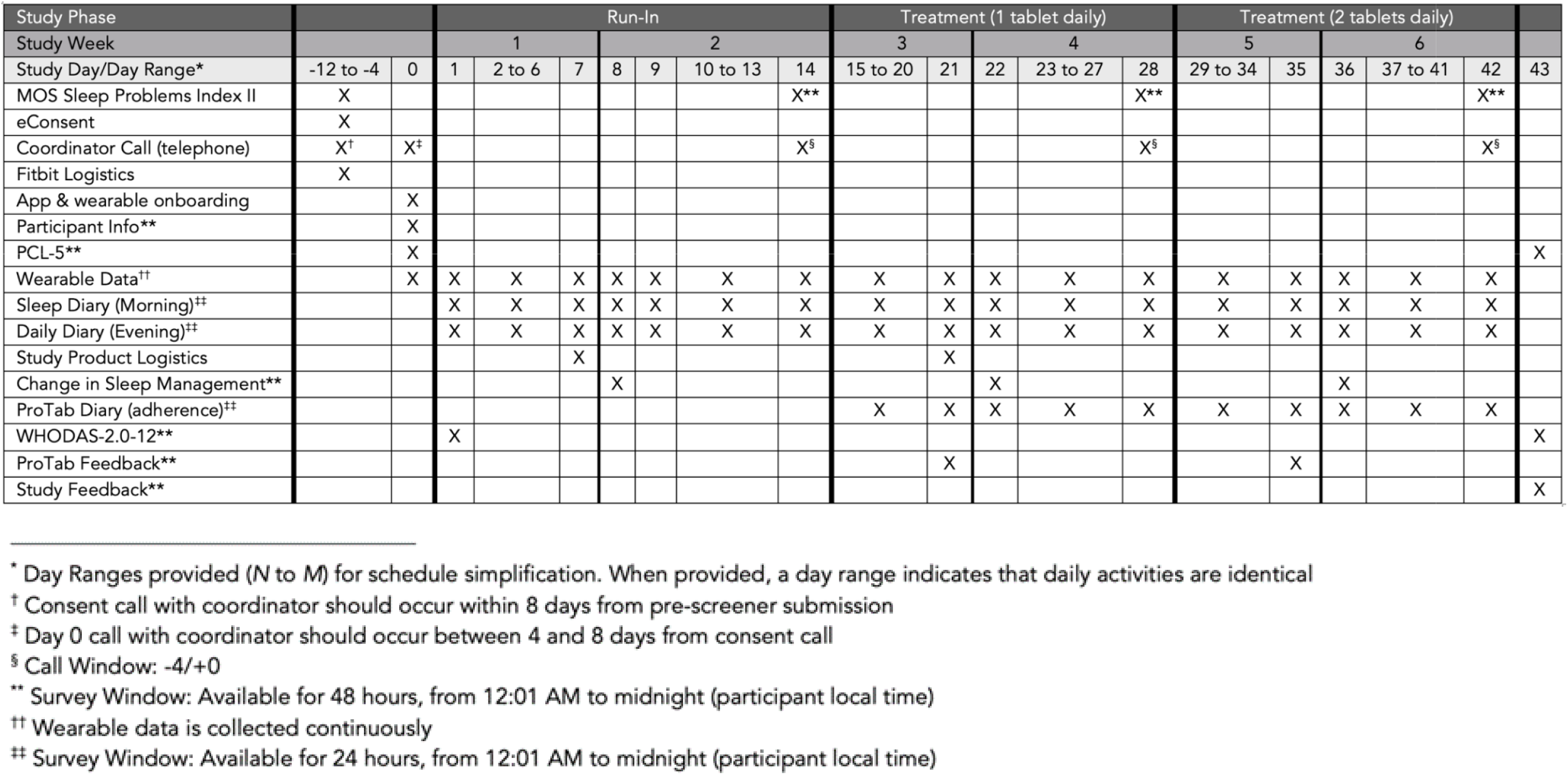
Veterans ECS21 Study Schedule of Assessments.

Participants will begin collecting biometric data using a Fitbit Inspire 2. After the two-week run-in period, participants will enter a four-week treatment period. For the first two weeks of treatment, participants will either take a daily dose of 25 mg CBG (CBG Protab™) (i.e., be assigned to the active treatment arm) or an identical-looking placebo formulation with no CBG (i.e., be assigned to the placebo control arm). For the last two weeks, participants in the active treatment arm will take two Protab™ tablets (50 mg CBG) and participants in the control arm will take two placebo tablets. Participants will continue answering regular questionnaires and collecting biometrics with the Fitbit (24-hours a day, seven-days a week wear time). During the treatment period, the nightly questionnaire will include dosing questions to monitor participants adherence. The study ends following the four-week treatment phase, with a total duration of approximately seven to eight weeks.

Participants will be assigned a dedicated Clinical Research Coordinator (CRC) who is trained on the unique, lived experiences of a Veteran. Participants will have a virtual, bi-monthly check-in call throughout the study to provide support, instructions, and answer questions. The study protocol and supporting documentation was approved by a central Institutional Review Board (Advarra; Pro00056526) and registered at clinicaltrials.gov (NCT05088018).

### Dosage and Dose Modifications

There is no known ‘therapeutic’ dose for CBG. CBG products are widely commercially available in a variety of formats (gummies, isolate powders, oils, tinctures, sprays, and more), both with or without other cannabinoids. Since anecdotal reports indicate a positive impact using commercially available CBG, we selected a target dosage range that mirrored commonly available dosages with an upper range of 50 mg daily to avoid tolerability issues. Studies of similar non-psychoactive, orally-administered cannabinoids such as CBD reported dosages of 400 mg and 800 mg of CBD were well-tolerated.^43-45^ As CBG has no psychoactive effect, the upper threshold of dosing is not restricted due to potential impairment.

We anticipate a build up of CBG over time, like with other highly lipophilic cannabinoids. For example, the half-life of CBD after oral administration may be 2-5 days.^44^ By escalating the dosage mid-way through the treatment phase (i.e., after two weeks), we hope to better evaluate the effective dose of CBG for the improvement of sleep and study any potential dose-dependent effects.

Participants will be asked to self-administer a daily dose according to the dosage instructions (1 tablet or 2 tablets, depending on where they are in the study schedule) and will report whether they dosed, the time of dosing, and how many tablets they took through the evening daily questionnaire in the study application. Participants will be asked to take their study product any time between when they wake up but no more than three hours before bed.

### Safety Monitoring

The safety of participants will be measured and monitored throughout the study by assessing subjective study product effects, psychological distress and adverse events (AEs). As C.R. Emerson is the study’s principal investigator but is not a medical doctor, we have engaged a clinician so as to delegate safety monitoring responsibilities during the study.

We recognize that questionnaires like the PCL-5 ask individuals to discuss a traumatic experience. We designed the study application to trigger an alert to an on-call clinician when specific responses might indicate the participant is experiencing a current and severe crisis. The on-call clinician is a psychiatrist with experience treating persons who have experienced trauma. They will reach out to the participants to recommend sources of emergency or non-emergency support, based on their assessment of the participant at the time of the call.

### Study Application

Study participants will use a web-based electronic data capture system for the collection of clinical trial data within electronic case report forms. This application hosted by Curebase collects all study questionnaire data and any adverse events reported during the study. It also displays each participant’s study calendar for their check-in calls. Email and text message reminders of upcoming check-in calls and questionnaire due dates will be provided occasionally throughout the study. The study application requires internet access in order to submit any data, but can be accessed on either a smartphone, tablet, or a laptop.

### Wearable Device and Objective Measures

The Fitbit Inspire 2 is a lightweight, wrist-worn activity tracking sensor that includes a three-axis accelerometer and an infrared optical pulse sensor. The battery life can be up to 10 days with a single charge (www.fitbit.com).^46^ There is a one-time set-up process where the participant will link the sensor to the Fitbit smartphone app and then authorize it to share its data with the study application. Throughout the study, the participant will only need to open the Fitbit smartphone app to sync data over Bluetooth. Participants are instructed to wear the Fitbit at all times, (i.e., twenty-four hours a day, seven days a week), with guidance to only remove the sensor and charge it while they shower.

The data streams of highest interest to our analysis include:

- Sleep tracking: Sleep efficiency; time spent in light, deep, and REM sleep stages
- Activity tracking: Steps (collected in 1 second intervals); time spent in ‘fairly active’, ‘lightly active’, ‘sedentary’, and ‘very active’ activity stages
- Heart rate: Heart rate over time (collected in 1 second intervals) and resting heart rate

### Statistical Analysis Plan

Our primary endpoint will evaluate the change, per participant, between the MOS-SS SPI-II score at the end of the study (after four weeks of treatment) and the score obtained during the run-in phase (prior to treatment). The first exploratory objective for characterizing the effective dose will be evaluated likewise, except by comparing the MOS-SS SPI-II score after two weeks of treatment at 25 mg daily CBG to the MOS-SS SPI-II score after two weeks of treatment at 50 mg daily CBG.

MOS-SS SPI-II scores range from 0-100, with higher scores indicating a worse outcome. Its use in a similar study population (a RCT of non-Veterans adults exhibiting PTSD) reported baseline values of 57.0 (SD 16.8) and 60.0 (SD = 17.5) for each study arm (n=108 and n=101, respectively). The intervention, a holistic yoga program, resulted in a 12-point decrease while the “placebo”, a wellness lifestyle Mprogram, only resulted in a decrease of 4.6 points after 16 weeks of participation.^22^ For this study, we’ve estimated a 10-point difference as clinically meaningful and used a minimum score of 30 as an inclusion criterion; while this threshold does not constitute a diagnosed sleep issue, we felt it was an appropriate starting threshold by which to measure change for our study population.

Following the procedure outlined under the section “6.1 WHODAS 2.0 summary scores: Simple Scoring” from the World Health Organization,^25^ we will calculate the WHODAS-2.0-12 endpoints for each participant as our secondary endpoint, comparing the score at the beginning of the study to the end of the study. There is no clinically meaningful mean change defined in the literature as functional ability and score change vary widely by condition and intervention.^28^ In a large, non-interventional RCT in a Veteran population, participants reported an average score of 38.18 (SD 25.24, n=1,109) and found that the WHODAS-2.0 was an acceptable way to assess functional impairment among veterans seeking compensation for PTSD.^30^

Another study evaluating cognitive behavior therapy (CBT) in patients with stress and anxiety disorders reported baseline WHODAS-2.0–12 scores (using simple scoring) of 23.4 (SD, 7.9; range 12–51).^47^ That study indicated that the estimated minimal clinically important difference (MCID) ranged from 3 to 7 points.^47^ For the present study we considered a 5-point decrease as the minimum clinically meaningful average change.

We will calculate the remaining exploratory endpoints per participant as follows. Fitbit sensor data for calculating the endpoints for sleep duration and quality, activity duration, and heart rate will first be pre-processed and cleaned (i.e., device artifact removal, adjustment for wear-time and missing data, imputation of missing data if appropriate). We will then aggregate these by day, and will analyze these daily summary statistics (e.g., mean, median, range, standard deviation, interquartile range) as our exploratory outcomes. We will define the corresponding endpoint as pre–post.

For our main objectives, we want to statistically discern or detect a hypothesized true average difference (i.e., association or effect size) of size _δ_ or larger in continuous outcomes *Y* between 2 treatment groups. We will assume these outcomes vary with the same standard deviation (SD) _σ_*Y* in both groups. These two quantities are often combined as Cohen’s *d* (defined as *d* = _δ_*/*_σ_*Y*). We will also assume these outcomes are fairly normally distributed, allowing us to rely on Cohen’s *d* and the two-sample t-test in our calculations.

For our primary and secondary endpoints, we will use a Bonferroni correction that equally splits the familywise _α_ in 2 (i.e., each test’s _α_ is equal to _α_/2). For each main-objective endpoint, we will estimate the average treatment effect after all four weeks of treatment, specified as the mean difference in endpoints under active treatment versus placebo control. We will also conduct a t-test to quantify the amount of statistical evidence (i.e., statistical significance) for this mean difference not being equal to zero; i.e., to quantify how statistically discernible (i.e., statistically significant) the mean difference is from zero given our sample.

For exploratory objective 1, we are most interested in how different the cumulative average treatment effect on MOS-SS SPI-II score is between the different dosing i.e., 25 mg CBG daily after the first two weeks of treatment, versus the overall change after all four weeks of treatment (with 25 mg CBG daily for 2 weeks and then 50mg CBG daily for 2 weeks). Our goal for exploratory objectives 2–4 will be to generate scientific hypotheses for future study. That is, we will first visualize and otherwise explore the structure of the data. Based on what we find, we will decide how to define treatment effects of interest. These post hoc hypotheses will help us define future hypotheses for appropriately powered confirmatory studies.

For each exploratory objective, we may fit post hoc statistical models or conduct statistical hypothesis tests that also account for baseline measurements. Our goal in fitting these models will be to characterize how each treatment effect might vary after accounting for particular levels of these baseline characteristics, or across such levels (e.g., the treatment effect on MOS-SS SPI-II might be higher or lower for older participants who do not use tobacco, modeled by interacting treatment with tobacco use).

### Data Management

The Curebase application is 21 CFR Part 11 compliant and exists in a secure cloud architecture which is hosted on Amazon Web Services (AWS). The Curebase server collects data from the participant’s Fitbit wearable sensors. Authentication and authorization are managed through OAUTH2 over HTTPS from the Fitbit website.

Data is retrieved directly from vendor servers through their public APIs and sent to Curebase, where all processing is conducted over an encrypted TLS 1.2 HTTPS connection.

### Study Partners

Metta Medical Inc (doing business as LEVEL) is an effects-based hemp and cannabis company. Metta’s LEVEL products (https://levelexperience.com/) give customers cannabinoid and formulation options to provide a consistent, scientifically developed, and outcome-focused experience. LEVEL products are legally marketed and distributed within the within the state of California. Metta Medical is the study sponsor and provides the CBG and placebo Protab™ study interventional product.

The Veterans Cannabis Group (https://veteranscannabisgroup.com/), the study co-sponsor, is a non-profit, 501(c)(3) advocacy group of Veterans for Veterans who use medical cannabis. VCG provides education, safe access, information on VHA resources and benefits, employment, and networking opportunities in the cannabis industry, and supports veteran owned cannabis businesses from cultivation to retail. Additional recruitment partners include Operation EVAC (Educating Veterans About Cannabis, http://www.opevac.org/) and the Santa Cruz Veterans Alliance (https://www.scva.us/). Additional Veterans groups may participate in distributing study recruitment materials, though all groups and study participants will be restricted to the state of California.

Curebase is responsible for deploying the study application, secure data hosting, and provides the remote clinical study coordinators supporting the participant.

## Discussion

The Veteran population has experienced a six-fold increase in sleep disorders from 2000 to 2010, and is more likely to be impacted by sleep-related issues than the general population.^1^ New approaches to treat and support Veterans are critically needed. Though commercially available and anecdotally reported to improve sleep, there are no controlled human studies of CBG to evaluate its efficacy. This pilot study aims to gather preliminary evidence of the impact of CBG on sleep quality.

### Mitigating Study Design Challenges

In the Introduction, we outlined four challenges to overcome in this study’s design.

#### Challenge #1: Recruiting and engaging a skeptical Veteran community

We need to engage a community with an understandable mistrust of sharing sensitive personal information. We hosted “Participant Panels” to hear the voice of this community and get their input on study design. We are extremely grateful to the Veterans that volunteered their time and their openness in sharing their experiences. We showed panel participants a few questions from the PCL-5 to gauge how willing they would be to answer sensitive questions on a mobile device. We learned how familiar they are with mental health questionnaires and their reluctance to answer these questions honestly. Below are direct responses by Veterans who participated in the Participant Panels:

- “*I answered these two days ago in my therapy session. It feels sterile. Not all of this applies to me, so I’m just going to do my best to answer something*.”
- “*The problem with these questions is that if you answer them, you can get 5150’d. They are just trying to figure out if you’re a problem, they aren’t asking if you have a hole in your center*.”
- “*Am I going to incriminate myself? I would not answer that question. I’m not saying I wouldn’t do it, but would I answer it honestly? There are some things I’m just never going to write down anywhere*.”
- [When asked these questions frequently] “*You start to judge yourself, like ‘What do they want me to be saying?*’”

While we couldn’t modify the validated questionnaires, this feedback helped us craft the following introduction we used before each questionnaire:

- “These questions will help us learn more about you and your personal experiences. There are no “right” or “wrong” answers because everyone’s experience is unique to them. If you feel stuck or you are not sure how to answer a question, just do the best you can. Remember: How you respond to these questions will not go on your medical record or be reported to the VHA, so please answer honestly. This information will be used only for this research study.”

We also heard how Veteran’s groups gave them back the sense of community they had in the military. We learned how challenging it was to feel “heard” and develop consistent relationships when seeking care:

- “*Getting a regular appointment is almost impossible*”
- “*Every time I switch doctors, they want to touch my meds*”
- “*It’s hard to relate to doctors that don’t know military culture*”

This validated our intention to recruit through Veterans advocacy organizations. We asked the Veterans Cannabis Group to be our study co-sponsor and for permission to use their logo on our recruitment materials. This provided Veterans with representation from their community during recruitment and provided our study team with a consistent and integrated source of feedback through study design. We emphasized the study sponsor’s Veteran status (e.g., “By Veterans For Veterans”) and how the mission of this study is to support Veterans issues.

#### Challenge #2: Recruiting and retaining a representative study population

Early on, we envisioned that a decentralized study design might lower access-related barriers to clinical research for our target population, as nearly 70% of potential clinical trial participants live more than two hours away from a study center.^48^ In In addition to recruitment challenges, we also needed to maintain adherence and retention for at least six weeks. With the relative novelty of fully remote research studies, the research community is still learning how to engage decentralized study participants.

For this study, we wanted each study participant to meet with the same CRC for their study check-in’s. This models the “Engagement Specialist” methodology recently published in the BUMP study by S. Goodday, et. al.^49^ We prepared training for these CRCs using the insights we obtained during the participant panels (**Figure 2: Cultural Competency Guidance developed for CRC Training**) to ensure they could understand the Veterans unique experience. Lastly, we designed recurring check-in questions to foster a relationship and provide support (**Supplementary Table 2**).

**Figure 2:**
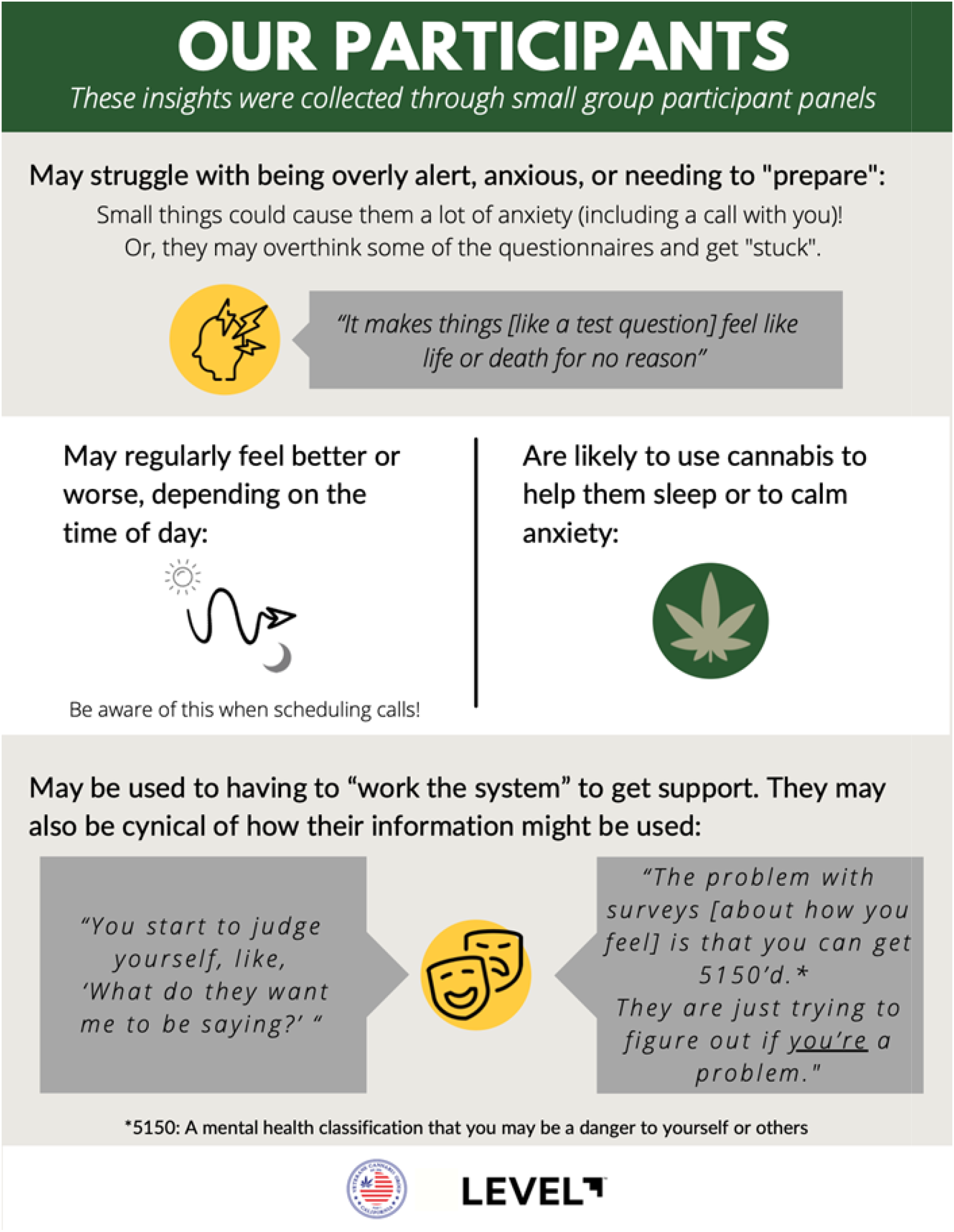
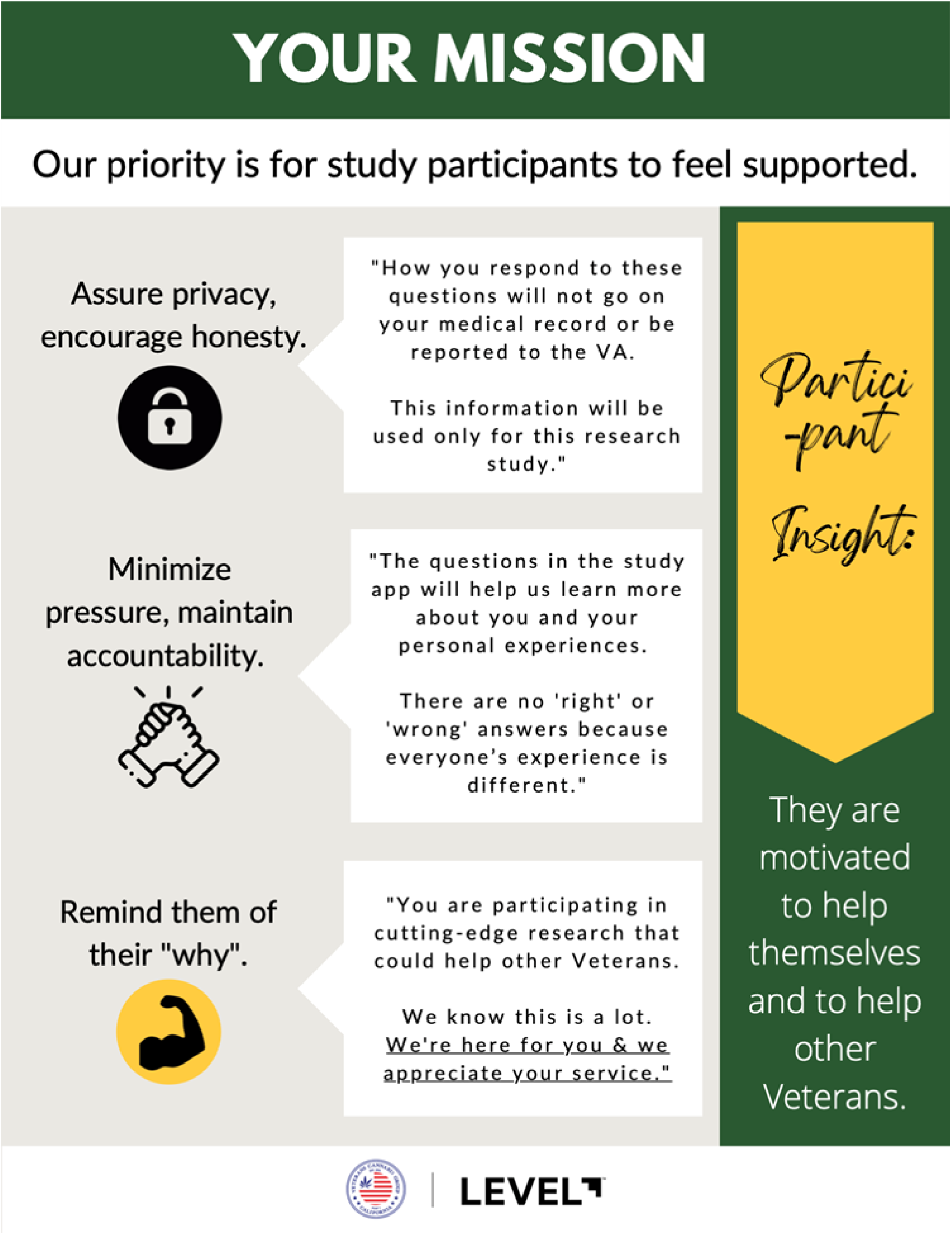
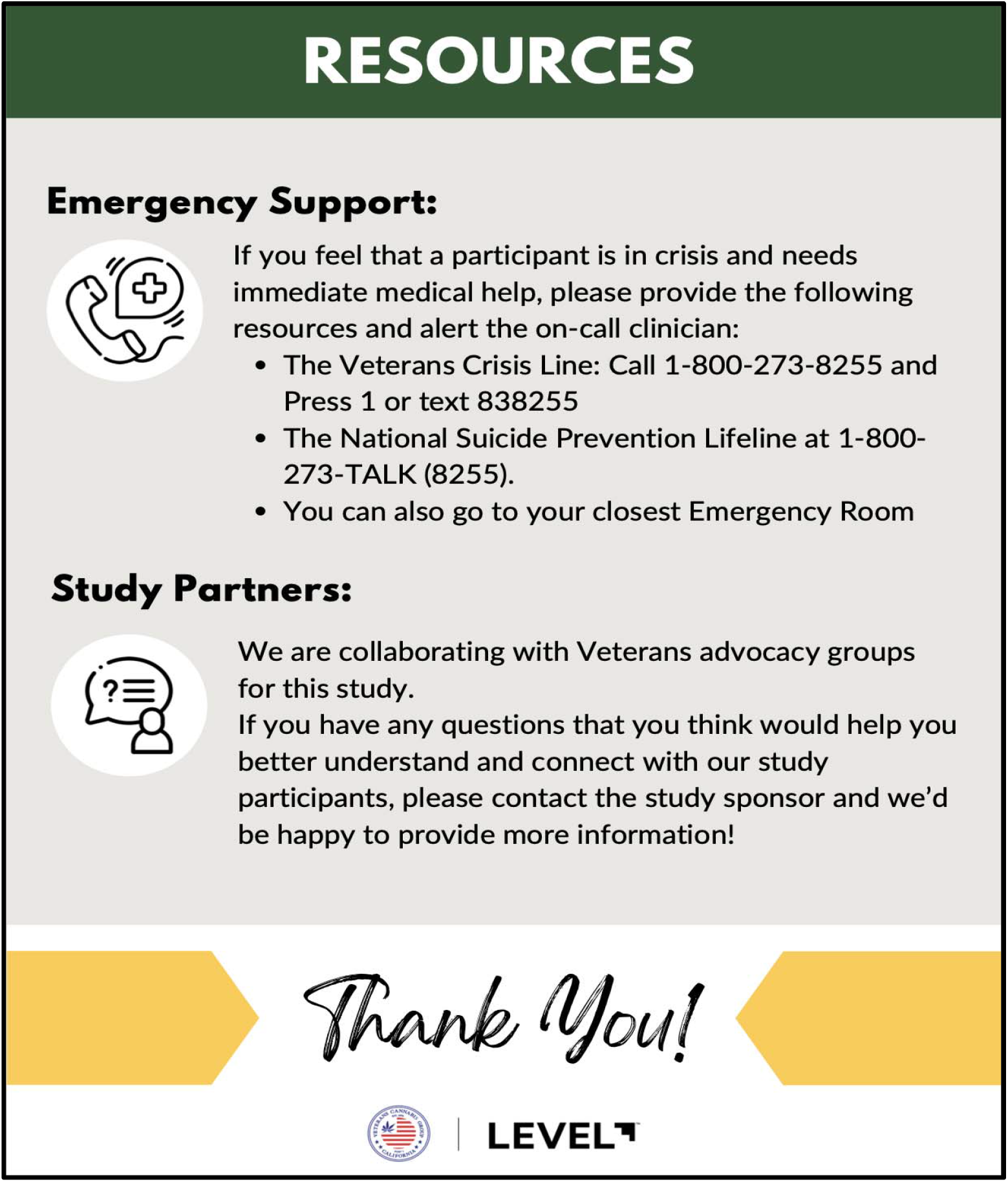
Cultural Competency Guidance developed for CRC Training (Excerpt)

#### Challenge #3: Minimizing or avoiding common confounders in cannabis research

Most cannabis studies use an inhalable method of administration and use the cannabis flower, which contains variable quantities of hundreds of active compounds. There are additional benefits for using a non-psychoactive cannabinoid for this study, as cannabis with a psychoactive effect is difficult to placebo control and presented more safety-related concerns.

LEVEL’s CBG Protab™ is a swallowable tablet that allows us to control dosage, dose reliability and generate a realistic placebo. The placebo study product will be manufactured and packaged identically to the active study product, but the placebo Protab™ will not contain any CBG. Each batch of the investigational product will be evaluated by an independent laboratory to ensure that the cannabinoid profile was within established limits and with no _Δ_9-THC element.

We also selected a pre–post study design to evaluate within-individual changes before and after the introduction of CBG. Furthermore, the trial design is randomized, placebo-controlled, and triple-blinded (i.e., the participant; data collection, management, and analysis personnel; and the principal investigator will remain blinded).

#### Challenge #4: Effectively measuring a change in in sleep quality: Endpoint & Digital Health Technology (DHT) design

As previously noted, we desired a sleep PRO as our primary endpoint. We used traditional methods to select an appropriate questionnaire (e.g., evaluating content validity within a similar population, reliability, and usability characteristics such as recall period, availability, and cost

We wanted an objective digital measurement of sleep to use as a comparator to the subjectively reported outcome. There are a number of sleep biometrics to choose from. We considered nightly sleep efficiency (calculated by dividing the amount of time spent asleep by the total amount of time in bed) and duration of REM sleep as one of the most relevant digital outcomes for this study. We ultimately chose a Fitbit after using the Digital Medicine Society’s “V3” framework to guide our fit-for-purpose device selection. This framework relies on verification, analytical validation, and clinical validation,^40^ as well as considering traditional constraints such as cost, accessibility, usability, and other parameters. We knew that a Fitbit device performed adequately in healthy populations when compared to PSG (analytical validation),^50,51^ and had been clinically evaluated in populations with healthy^52^ and interrupted sleep.^53-55^

While we are aware of the limitations of a consumer-grade device when compared to a research-grade device, it was acceptable given the exploratory nature of the biometric endpoints. Further, we preferred a sensor that offered the ability to return usable health data back to the study population. Study participants would be able to view activity (calories, steps, miles), sleep data (sleep and wake times, total duration, a proprietary sleep “score”), as well as other data. Veterans have previously self-reported that one motivation behind wearing a Fitbit in a research setting was an increased self-awareness.^56^ Since we had a fully remote study design, we wanted to reinforce engagement and value however possible, and this seemed like an additional way we could honor Veterans’ preferences. We selected the Fitbit Inspire 2 for use in this study. The sleep algorithm and hardware were not expected to deviate greatly from prior studies with a similar multisensory approach and algorithm.

### Known Limitations

One limitation to this study is that it will only be offered in English. The VHA does not track English language proficiency, so it is unknown how many Veterans have limited English skills. The Migration Policy Institute (www.migrationpolicy.org) reported that only 2.6% of a population of Veterans that were either foreign-born or children of immigrants report limited English skills.^57^ If we assume that data is also representative of this enrolled cohort, that would result in only 2-3 study participants requiring non-English support on the scale of this study.

The study application for answering questionnaires could be used on a mobile phone or computer, but participants needed to have their own smartphone in order to connect the Fitbit device through the Fitbit app. The authors are aware of the ‘digital divide’ and how decentralized studies can be overly reliant on internet access and smartphone ownership.^58^ While in California 91% of households reported high-speed internet access in 2021, the adoption rate is still skewed higher for high-income households.^59^

We plan to account for these limitations in the subsequent, larger controlled trials that will be guided by our findings.

## Data Availability

All data produced in the present study are available upon reasonable request to the authors

## Acknowledgements

This study was funded by Metta Medical (dba LEVEL). The authors sincerely appreciate our co-sponsor Veterans Cannabis Group and Jeremy Freitas for their time, effort, and feedback throughout the study design phase. We are particularly grateful for the Veterans that volunteered their time for our participant panels to share their experiences and guide our study design. The authors would also wish to thank Iain O’Neill (Freelance on behalf of nymbly) for medical writing support.

## Author contributions

C.E.W. and C.R.E. conceptualized and designed the study. E.J.D. developed the Statistical Analysis Plan. C.E.W. wrote the first draft of the manuscript and all authors contributed to subsequent manuscript revisions.

## Table 1. Inclusion and Exclusion Criteria

### Inclusion Criteria

- Veteran status
- MOS Sleep Problems Index II > 30
- California resident
- Age 21 or over
- Participants must own their own device to use for the study. Devices must meet the following criteria:
  - Apple iOS 12.2 or higher
  - Android OS 7.0 or higher
- Participants must be comfortable reading study instructions in English and communicating with study team in English
- Be willing to commit to study dosing, completing evaluation instruments, and following study protocol activities.
- If female and of childbearing potential, agree to use an effective form of birth control during study participation; defined as those which result in a low failure rate (i.e., less than 1% per year) when used consistently and correctly such as implants, injectables, oral contraceptives, IUDs, or a vasectomized partner.
- If using sleep medications, medication and dosage have not been changed in the past month and will remain unchanged for the duration of the study
- If using other psychotropic medications, medication and dosage have not been changed in the past 2 months, and will remain unchanged for the duration of the study.
- If diagnosed with sleep apnea (participant reported), participant must be currently using a CPAP with at least four weeks of prior CPAP use
- If prior observation that the participant has stopped breathing or observed choking/gasping during their sleep, participant must be currently using a CPAP with at least four weeks of prior CPAP use

### Exclusion Criteria

- Currently in a Cognitive Behavioral Therapy for Insomnia (CBTI) program
- Women who are currently pregnant, trying to become pregnant, or breastfeeding

## References

1. Alexander M, Ray MA, Hebert JR, et al. The National Veteran Sleep Disorder Study: Descriptive Epidemiology and Secular Trends, 2000-2010. Sleep 2016;39(7):1399–410. DOI: 10.5665/sleep.5972.

2. Folmer RL, Smith CJ, Boudreau EA, et al. Prevalence and management of sleep disorders in the Veterans Health Administration. Sleep Med Rev 2020;54:101358. DOI: 10.1016/j.smrv.2020.101358.

3. Williamson EM, Evans FJ. Cannabinoids in clinical practice. Drugs 2000;60(6):1303–14. DOI: 10.2165/00003495-200060060-00005.

4. Choi S, Huang BC, Gamaldo CE. Therapeutic Uses of Cannabis on Sleep Disorders and Related Conditions. J Clin Neurophysiol 2020;37(1):39–49. DOI: 10.1097/WNP.0000000000000617.

5. Cannabis Data Company (BDSA). Retail sales tracking. Share of Market cannabinoids by sales (Jan, 2021 through June, 2022). Available at: https://bdsa.com/products/retail-sales-tracking/. [Accessed 8 July 2022].

6. Metrik J, Bassett SS, Aston ER, Jackson KM, Borsari B. Medicinal versus Recreational Cannabis Use among Returning Veterans. Transl Issues Psychol Sci 2018;4(1):6–20. DOI: 10.1037/tps0000133.

7. Metrik J, Jackson K, Bassett SS, Zvolensky MJ, Seal K, Borsari B. The mediating roles of coping, sleep, and anxiety motives in cannabis use and problems among returning veterans with PTSD and MDD. Psychol Addict Behav 2016;30(7):743–754. DOI: 10.1037/adb0000210.

8. Babson KA, Sottile J, Morabito D. Cannabis, Cannabinoids, and Sleep: a Review of the Literature. Curr Psychiatry Rep 2017;19(4):23. DOI: 10.1007/s11920-017-0775-9.

9. Russo EB, Guy GW, Robson PJ. Cannabis, pain, and sleep: lessons from therapeutic clinical trials of Sativex, a cannabis-based medicine. Chem Biodivers 2007;4(8):1729–43. DOI: 10.1002/cbdv.200790150.

10. Shannon S, Lewis N, Lee H, Hughes S. Cannabidiol in Anxiety and Sleep: A Large Case Series. Perm J 2019;23:18–041. DOI: 10.7812/TPP/18-041.

11. Kuhathasan N, Dufort A, MacKillop J, Gottschalk R, Minuzzi L, Frey BN. The use of cannabinoids for sleep: A critical review on clinical trials. Exp Clin Psychopharmacol 2019;27(4):383–401. DOI: 10.1037/pha0000285.

12. Suraev AS, Marshall NS, Vandrey R, et al. Cannabinoid therapies in the management of sleep disorders: A systematic review of preclinical and clinical studies. Sleep Med Rev 2020;53:101339. DOI: 10.1016/j.smrv.2020.101339.

13. Nachnani R, Raup-Konsavage WM, Vrana KE. The Pharmacological Case for Cannabigerol. J Pharmacol Exp Ther 2021;376(2):204–212. DOI: 10.1124/jpet.120.000340.

14. Giovannitti JA, Jr., Thoms SM, Crawford JJ. Alpha-2 adrenergic receptor agonists: a review of current clinical applications. Anesth Prog 2015;62(1):31–9. DOI: 10.2344/0003-3006-62.1.31.

15. Naguy A. Clonidine Use in Psychiatry: Panacea or Panache. Pharmacology 2016;98(1-2):87–92. DOI: 10.1159/000446441.

16. Hays, R. D., Stewart, A. L. Sleep measures. In A. L. Stewart & J. E. Ware (eds.), Measuring functioning and well-being: The Medical Outcomes Study approach (pp. 235–259), Durham (NC): Duke University Press; 1992.

17. Hays RD, Martin SA, Sesti AM, Spritzer KL. Psychometric properties of the Medical Outcomes Study Sleep measure. Sleep Med 2005;6(1):41–4. DOI: 10.1016/j.sleep.2004.07.006.

18. Spritzer, K. L. & Hays, R. D. MOS Sleep Scale: A Manual for Use and Scoring, Version 1.0. Los Angeles, CA; 2003. Available at: https://labs.dgsom.ucla.edu/hays/files/view/docs/surveys/sleep/sleepman-112603.pdf. [Accessed 8 November 2022].

19. Nakamura Y, Lipschitz DL, Landward R, Kuhn R, West G. Two sessions of sleep-focused mind-body bridging improve self-reported symptoms of sleep and PTSD in veterans: A pilot randomized controlled trial. J Psychosom Res 2011;70(4):335–45. DOI: 10.1016/j.jpsychores.2010.09.007.

20. Nakamura Y. Evaluating a Novel Sleep-Focused Mind-body Rehabilitative Program for Veterans with mTBI and Other Polytrauma Symptoms: An RCT Study. U.S. Army Medical Research and Development Command. Utah University, Salt Lake City 2018.

21. Nakamura Y, Lipschitz DL, Donaldson GW, et al. Investigating Clinical Benefits of a Novel Sleep-Focused Mind-Body Program on Gulf War Illness Symptoms: A Randomized Controlled Trial. Psychosom Med 2017;79(6):706–718. DOI: 10.1097/PSY.0000000000000469.

22. Davis LW, Schmid AA, Daggy JK, et al. Symptoms improve after a yoga program designed for PTSD in a randomized controlled trial with veterans and civilians. Psychol Trauma 2020;12(8):904–912. DOI: 10.1037/tra0000564.

23. Nakamura Y, Lipschitz DL, Kuhn R, Kinney AY, Donaldson GW. Investigating efficacy of two brief mind-body intervention programs for managing sleep disturbance in cancer survivors: a pilot randomized controlled trial. J Cancer Surviv 2013;7(2):165–82. DOI: 10.1007/s11764-012-0252-8.

24. Digital Medicine Society (DiMe). The playbook. Available at: https://playbook.dimesociety.org/. [Accessed 8 November 2022].

25. World Health Organization. Measuring health and disability: manual for who disability assessment schedule WHODAS 2.0. Geneva: WHO Press; 2010. Available at: https://www.who.int/publications/i/item/measuring-health-and-disability-manual-for-who-disability-assessment-schedule-(-whodas-2.0). [Accessed 8 November 2022].

26. Ustun TB, Chatterji S, Kostanjsek N, et al. Developing the World Health Organization Disability Assessment Schedule 2.0. Bull World Health Organ 2010;88(11):815–23. DOI: 10.2471/BLT.09.067231.

27. Gold LH. DSM-5 and the assessment of functioning: the World Health Organization Disability Assessment Schedule 2.0 (WHODAS 2.0). J Am Acad Psychiatry Law 2014;42(2):173–81. (https://www.ncbi.nlm.nih.gov/pubmed/24986344).

28. Federici S, Bracalenti M, Meloni F, Luciano JV. World Health Organization disability assessment schedule 2.0: An international systematic review. Disabil Rehabil 2017;39(23):2347–2380. DOI: 10.1080/09638288.2016.1223177.

29. Marx BP, Wolf EJ, Cornette MM, et al. Using the WHODAS 2.0 to Assess Functioning Among Veterans Seeking Compensation for Posttraumatic Stress Disorder. Psychiatr Serv 2015;66(12):1312–7. DOI: 10.1176/appi.ps.201400400.

30. Bovin MJ, Meyer EC, Kimbrel NA, et al. Using the World Health Organization Disability Assessment Schedule 2.0 to assess disability in veterans with posttraumatic stress disorder. PLoS One 2019;14(8):e0220806. DOI: 10.1371/journal.pone.0220806.

31. Pearce M, Garcia L, Abbas A, et al. Association Between Physical Activity and Risk of Depression: A Systematic Review and Meta-analysis. JAMA Psychiatry 2022;79(6):550–559. DOI: 10.1001/jamapsychiatry.2022.0609.

32. Blevins CA, Weathers FW, Davis MT, Witte TK, Domino JL. The Posttraumatic Stress Disorder Checklist for DSM-5 (PCL-5): Development and Initial Psychometric Evaluation. J Trauma Stress 2015;28(6):489–98. DOI: 10.1002/jts.22059.

33. Weathers FW, Litz BT, Keane TM, Palmieri PA, Marx BP, Schnurr PP. The PTSD Checklist for DSM-5 (PCL-5) 2013. Scale available from the National Center for PTSD at http://www.ptsd.va.gov.

34. California Welfare and Institutions Code, ARTICLE 1. Detention of Mentally Disordered Persons for Evaluation and Treatment [5150 -5155]. Available at: https://leginfo.legislature.ca.gov/faces/codes_displaySection.xhtml?lawCode=WIC&sectionNum=5150 [Accessed 8 July 2022].

35. Cheney AM, Koenig CJ, Miller CJ, et al. Veteran-centered barriers to VA mental healthcare services use. BMC Health Serv Res 2018;18(1):591. DOI: 10.1186/s12913-018-3346-9.

36. US Department of Veterans Affairs (VA). Insomnia After Returning from Deployment. Available at: https://www.veteranshealthlibrary.va.gov/Encyclopedia/142,41442_VA. [Accessed 8 November 2022].

37. Nicosia FM, Kaul B, Totten AM, et al. Leveraging Telehealth to improve access to care: a qualitative evaluation of Veterans’ experience with the VA TeleSleep program. BMC Health Serv Res 2021;21(1):77. DOI: 10.1186/s12913-021-06080-5.

38. Pratap A, Neto EC, Snyder P, et al. Indicators of retention in remote digital health studies: a cross-study evaluation of 100,000 participants. NPJ Digit Med 2020;3:21. DOI: 10.1038/s41746-020-0224-8.

39. Colizzi M, Bhattacharyya S. Cannabis use and the development of tolerance: a systematic review of human evidence. Neurosci Biobehav Rev 2018;93:1–25. DOI: 10.1016/j.neubiorev.2018.07.014.

40. Goldsack JC, Coravos A, Bakker JP, et al. Verification, analytical validation, and clinical validation (V3): the foundation of determining fit-for-purpose for Biometric Monitoring Technologies (BioMeTs). NPJ Digit Med 2020;3:55. DOI: 10.1038/s41746-020-0260-4.

41. Manta C, Mahadevan N, Bakker J, et al. EVIDENCE Publication Checklist for Studies Evaluating Connected Sensor Technologies: Explanation and Elaboration. Digit Biomark 2021;5(2):127–147. DOI: 10.1159/000515835.

42. US Department of Veterans Affairs (VA). How Common is PTSD in Veterans? Available at: https://www.ptsd.va.gov/understand/common/common_veterans.asp. [Accessed 8 November 2022].

43. Kirkland AE, Fadus MC, Gruber SA, Gray KM, Wilens TE, Squeglia LM. A scoping review of the use of cannabidiol in psychiatric disorders. Psychiatry Res 2022;308:114347. DOI: 10.1016/j.psychres.2021.114347.

44. Millar SA, Stone NL, Yates AS, O’Sullivan SE. A Systematic Review on the Pharmacokinetics of Cannabidiol in Humans. Front Pharmacol 2018;9:1365. DOI: 10.3389/fphar.2018.01365.

45. Arnold JC, McCartney D, Suraev A, McGregor IS. The safety and efficacy of low oral doses of cannabidiol: An evaluation of the evidence. Clin Transl Sci 2023;16(1):10–30. DOI: 10.1111/cts.13425.

46. Fitbit Inspire 2. Available at: https://www.fitbit.com/global/us/products/trackers/inspire2. [Accessed 8 November 2022].

47. Axelsson E, Lindsater E, Ljotsson B, Andersson E, Hedman-Lagerlof E. The 12-item Self-Report World Health Organization Disability Assessment Schedule (WHODAS) 2.0 Administered Via the Internet to Individuals With Anxiety and Stress Disorders: A Psychometric Investigation Based on Data From Two Clinical Trials. JMIR Ment Health 2017;4(4):e58. DOI: 10.2196/mental.7497.

48. Shore C, Khandekar E, Alper J. Virtual Clinical Trials: Challenges and Opportunities: Proceedings of a Workshop. Washington (DC) 2019. DOI: 10.17226/25502.

49. Goodday SM, Karlin E, Brooks A, et al. Better Understanding of the Metamorphosis of Pregnancy (BUMP): protocol for a digital feasibility study in women from preconception to postpartum. NPJ Digit Med 2022;5(1):40. DOI: 10.1038/s41746-022-00579-9.

50. Haghayegh S, Khoshnevis S, Smolensky MH, Diller KR, Castriotta RJ. Accuracy of Wristband Fitbit Models in Assessing Sleep: Systematic Review and Meta-Analysis. J Med Internet Res 2019;21(11):e16273. DOI: 10.2196/16273.

51. Beattie Z, Oyang Y, Statan A, et al. Estimation of sleep stages in a healthy adult population from optical plethysmography and accelerometer signals. Physiol Meas 2017;38(11):1968–1979. DOI: 10.1088/1361-6579/aa9047.

52. de Zambotti M, Cellini N, Goldstone A, Colrain IM, Baker FC. Wearable Sleep Technology in Clinical and Research Settings. Med Sci Sports Exerc 2019;51(7):1538–1557. DOI: 10.1249/MSS.0000000000001947.

53. Hamill K, Jumabhoy R, Kahawage P, de Zambotti M, Walters EM, Drummond SPA. Validity, potential clinical utility and comparison of a consumer activity tracker and a research-grade activity tracker in insomnia disorder II: Outside the laboratory. J Sleep Res 2020;29(1):e12944. DOI: 10.1111/jsr.12944.

54. Wulterkens BM, Fonseca P, Hermans LWA, et al. It is All in the Wrist: Wearable Sleep Staging in a Clinical Population versus Reference Polysomnography. Nat Sci Sleep 2021;13:885–897. DOI: 10.2147/NSS.S306808.

55. Stucky B, Clark I, Azza Y, et al. Validation of Fitbit Charge 2 Sleep and Heart Rate Estimates Against Polysomnographic Measures in Shift Workers: Naturalistic Study. J Med Internet Res 2021;23(10):e26476. DOI: 10.2196/26476.

56. Ng A, Reddy M, Zalta AK, Schueller SM. Veterans’ Perspectives on Fitbit Use in Treatment for Post-Traumatic Stress Disorder: An Interview Study. JMIR Ment Health 2018;5(2):e10415. DOI: 10.2196/10415.

57. Migration Policy Institute Immigrant Veterans in the United States Available at: https://www.migrationpolicyorg/article/immigrant-veterans-united-states-2018 [Accessed 8 November 2022].

58. Goodson N, Wicks P, Morgan J, Hashem L, Callinan S, Reites J. Opportunities and counterintuitive challenges for decentralized clinical trials to broaden participant inclusion. NPJ Digit Med 2022;5(1):58. DOI: 10.1038/s41746-022-00603-y.

59. California Emerging Technology Fund (CETF). CETF-USC Statewide Broadband Adoption Survey. 2021. Available at: https://www.cetfund.org/wp-content/uploads/2021/03/Statewide-Survey-on-Broadband-Adoption-CETF-Report.pdf. [Accessed 8 November 2022].

