## Supplementary Material for "The Veterans ECS21 (Exploring Cannabigerol for Sleep) Study: A protocol for the first randomized clinical trial of Cannabigerol (CBG), a decentralized study designed for U.S. Veterans in California"

***Corresponding author**

### **Supplementary Table 1: Share of Market (SOM) cannabinoids by sales (Jan, 2021 through June, 2022) - BDSA^1^**

| **Cannabinoid** | **Share of Market** |
| --- | --- |
| THC | 94.56% |
| CBD | 4.46% |
| CBN | 1.34% |
| CBG | 0.26% |
| THCa | 0.14% |
| Delta 8 | 0.08% |
| CBDa | 0.04% |
| THCV | 0.04% |
| CBC | 0.00% |

SOM is calculated by searching for the cannabinoid listed within the product name. This leads to the total SOM of 100.93%, representing that this is not a perfect method as not all cannabinoids used in the product may be listed by BDSA

### **Supplementary Table 2: Participant Panel Excerpt**

*“As part of this study, we’d like to include a survey that asks about PTSD symptoms in detail. You might have answered a survey like this before at the VA or a doctor’s office. It’s important that we ask the questions in the exact same way as a doctor would. This survey has 20 questions and we think it should take about 10 minutes to complete.*

*Can you take a moment to read some of the questions?”*

| **Question** | **Response Options** |
| --- | --- |
| In the past month, how much were you bothered by:  Repeated, disturbing, and unwanted memories of the stressful experience? | Not at all  A little bit  Moderately  Quite a bit  Extremely |
| In the past month, how much were you bothered by:  Repeated, disturbing dreams of the stressful experience? | Not at all  A little bit  Moderately  Quite a bit  Extremely |
| In the past month, how much were you bothered by:  Suddenly feeling or acting as if the stressful experience were actually happening again (as if you were actually back there reliving it)? | Not at all  A little bit  Moderately  Quite a bit  Extremely |

*“Would you be willing to answer these questions on a smartphone app?”*

*“What, if anything, would make answering these questions easier?”*

### **Supplementary Table 3: Study Instruments**

#### **Pre-Screener Questionnaire**

| **Pre-Screener:**  *Approximate time to complete: 8-12 min* | | | |
| --- | --- | --- | --- |
| # | Question | Response Options | Notes |
| N/A | Let’s get started! We will provide you with more details so you can decide if this study is right for you.  These questions will help us learn more about you and your personal experiences. There are no “right” or “wrong” answers because everyone’s experience is unique to them. If you feel stuck or you are not sure how to answer a question, just do the best you can.  **Remember: How you respond to these questions will not go on your medical record or be reported to the VA, so please answer honestly. This information will be used only for this research study.** | N/A | Introduction |
| 1 | Are you 21 years of age or older? | Yes  No | Single-select |
| 2 | Do you own your own smartphone that you can use for this study and do you have a reliable connection to the internet at your home? | Yes  No | Single-select |
| 3 | How comfortable do you feel reading English on your phone? | Comfortable  Uncomfortable | Single-select |
| 4 | How comfortable do you feel speaking in English with others? | Comfortable  Uncomfortable | Single-select |
| 5 | Are you a Veteran of the U.S. military? | Yes  No | Single-select |
| 6 | Are you a resident of the state of California and will you be living in California for the next 7-8 weeks? | Yes  No | Single-select |
| 7 | Are you regularly taking prescribed or over-the-counter medications or supplements for sleep?  For example: Ambien, Zolpidem, Lunesta, benzodiazepines, unisom, diphenhydramine, doxylamine, melatonin, valerian, etc? | Yes  No | Single-select |
| 7a | In the last 2 months, have you started this sleep aid, changed what sleep aid you are taking, or changed your dosage? | Yes  No  I’m not sure | **Only presented if they select “Yes” to Question 7**  Single-select |
| 7b | If you participate in the study, do you agree to stay on this sleep aid for the next 6-8 weeks? | Yes  No | **Only presented if they select “No” to Question 7a**  Single-select |
| 8 | Within the last two months, have you stopped taking prescribed or over-the-counter medications or supplements for sleep?  For example: Ambien, Zolpidem, Lunesta, benzodiazepines, unisom, diphenhydramine, doxylamine, melatonin, valerian, etc? | Yes  No | Single-select |
| 9 | *[Removed - Amendment v1.2]* |  |  |
| 10 | Has anyone observed you stop breathing or choking/gasping during your sleep? | Yes  No | Single-select |
| 10a | Do you currently use a positive airway pressure device, like a CPAP, for sleep apnea? | Yes  No | **Only presented if they select “Yes” to Question 10**  Single-select |
| 10b | Have you been using this positive airway pressure device for the last four weeks (or more)? | Yes  No | **Only presented if they select “Yes” to Question 10a**  Single-select |
| 11 | Have you been diagnosed by a doctor with sleep apnea? | Yes  No | Single-select |
| 11a | Do you currently use a positive airway pressure device, like a CPAP, for sleep apnea? | Yes  No | **Only presented if they select “Yes” to Question 11**  Single-select |
| 11b | Have you been using this positive airway pressure device for the last four weeks (or more)? | Yes  No | **Only presented if they select “Yes” to Question 10a**  Single-select |
| 12 | Are you currently prescribed medications to manage any mental health symptoms (for example: Zoloft, Paxil, Lexapro, Cymbalta, Prozac, Celexa, Risperdal, Abilify, etc)? | Yes  No | Single-select |
| 12a | In the last 2 months, have you started this medication, changed which medication you are taking or changed your dosage? | Yes  No  I’m not sure | **Only presented if they select “Yes” to Question 12**  Single-select |
| 12b | If you participate in the study, do you agree to stay on this medication for the next 6-8 weeks? | Yes  No | **Only presented if they select “No” to Question 12a**  Single-select |
| 13 | Within the last two months, have you stopped taking medications to manage any mental health symptoms (for example: Zoloft, Paxil, etc)? | Yes  No | Single-select |
| 14 | Are you currently in a therapy program specifically for insomnia, such as CBTI (Cognitive Behavioral Therapy for Insomnia)? | Yes  No | Single-select |
| 15 | What is your sex as assigned at birth? | Male  Female | Single-select |
| 15a | Are you currently pregnant, trying to become pregnant, or breastfeeding? | Yes  No | **Only presented if they select “Female” to Question 15**  Single-select |
| 16 | Are you willing to spend approximately 5 minutes a day answering surveys for 6 weeks? | Yes  No | Single-select |
| 17 | Are you willing to wear a FitBit during the study?  (The FitBit will be provided to you and you can keep it after the study is complete) | Yes  No | Single-select |
| 18 | Do you regularly use cannabis products that are highly enriched in cannabigerol (CBG)?  For example: A CBG isolate, oil, or 1:1 tincture of CBG:CBD would be considered highly enriched. Cannabis flower is not considered highly enriched in CBG. | Yes  No | Single-select |
| 18a | Can you tell us more about what you’re taking that’s highly enriched in cannabigerol (CBG)? For example, the brand name and amount of CBG. | Free text | **Only presented if they select “Yes” to Question 18** |
| *N/A* | *If qualified so far, participants should proceed to the MOS Sleep Problems Index II. Participants must score >30 to be fully qualified for the study.* | *N/A* | *Not participant-facing content* |

#### **Participant information Questionnaire**

| **Participant Info:**  *Approximate time to complete: 5-9 min* | | | |
| --- | --- | --- | --- |
| # | Question | Response Options | Notes |
| N/A | These questions will help us learn more about you and your personal experiences. There are no “right” or “wrong” answers because everyone’s experience is unique to them. If you feel stuck or you are not sure how to answer a question, just do the best you can.  **Remember: How you respond to these questions will not go on your medical record or be reported to the VA, so please answer honestly. This information will be used only for this research study. Please see your Informed Consent Form for more information on how we keep your information private.** | N/A | Introduction |
| 1 | How would you describe yourself? | American Indian or Alaska Native  Asian  Black or African American  Native Hawaiian or Other Pacific Islander  White  Other (please specify) | Multi-select |
| 2 | What is your date of birth? | MMM-DD-YYYY | Date |
| 3 | What is your current weight (in pounds)? | NNN | Integer |
| 4 | What is your current height? | NN Feet, NN inches | Integer |
| 5 | Do you regularly smoke tobacco? | Yes  No | Single-select |
| 6 | Do you regularly use cannabis products? | Yes  No | Single-select |
| 6a | What type of cannabis product(s) do you regularly use? | THC  CBD  Both THC and CBD  I don’t know | **Only presented if they select “Yes” to Question 6** |
| 6b | How often do you normally use cannabis? | Multiple times a day  About once a day  About every other day  Once or twice a week  Once or twice a month  Never/Rarely | **Only presented if they select “Yes” to Question 6 and following Question 6a** |
| 7 | Please select any sources of support you regularly use to help you sleep: | Sleep therapy  Meditation  Exercise  Other (text response) | Multi-select |
| 8 | How long ago did you start experiencing sleep-related issues? | Less than a year ago  Around 1-2 years ago  Around 2-5 years ago  Around 5-10 years ago  More than ten years ago  I’m not sure | Single-select |
| 9 | Do you have times of the day when you *regularly* feel better or worse? | Yes  No  I’m not sure | Single-select |
| 9a | During what part of the day do you typically feel the best? (Select all that apply) | Morning  Mid-day  Afternoon  Evening  Night-time  I’m not sure | **Only presented if they select “Yes” to Question 9** |
| 9b | During what part of the day do you typically feel the worst? (Select all that apply) | Morning  Mid-day  Afternoon  Evening  Night-time  I’m not sure | **Only presented if they select “Yes” to Question 9** |

#### **MOS-SS SPI-II**

| **MOS Sleep Problems Index II**  Approximate time to complete: 2-3 min | | | |
| --- | --- | --- | --- |
| # | Question | Response Options | Notes |
| N/A | These questions will help us learn more about you and your sleep. We need to ask these questions the same way a doctor would, so you may have seen these questions before at the VA.  It’s ok if some of these questions are hard for you to answer. There are no “right” or “wrong” answers because everyone’s experience is unique to them. If you feel stuck or you are not sure how to answer a question, just do the best you can.  Remember: How you respond to these questions will not go on your medical record or be reported to the VA, so please answer honestly. This information will be used only for this research study. Please see your Informed Consent Form for more information on how we keep your information private. | N/A | Introduction |
| N/A | Sleep Scale from the Medical Outcomes Study  This survey was reprinted with permission from the RAND Corporation. Copyright © the RAND Corporation. RAND's permission to reproduce the survey is not an endorsement of the products, services, or other uses in which the survey appears or is applied. | N/A | Introduction |
| 1 | How long did it usually take for you to fall asleep during the past 2 weeks? | 0-15 minutes  16-30 minutes  31-45 minutes  46-60 minutes  More than 60 minutes | Single-select |
| 2 | On the average, how many hours did you sleep each night during the past 2 weeks?  (Enter the number of hours per night) | NN | Integer from 0-24 |
| 3 | How often during the past 2 weeks did you feel that your sleep was not quiet (moving restlessly, feeling tense, speaking, etc., while sleeping?) | All of the Time  Most of the Time  A Good Bit of the Time  Some of the Time  A Little of the Time  None of the Time | Single-select |
| 4 | How often during the past 2 weeks did you get enough sleep to feel rested upon waking in the morning? | All of the Time  Most of the Time  A Good Bit of the Time  Some of the Time  A Little of the Time  None of the Time | Single-select |
| 5 | How often during the past 2 weeks did you awaken short of breath or with a headache? | All of the Time  Most of the Time  A Good Bit of the Time  Some of the Time  A Little of the Time  None of the Time | Single-select |
| 6 | How often during the past 2 weeks did you feel drowsy or sleepy during the day? | All of the Time  Most of the Time  A Good Bit of the Time  Some of the Time  A Little of the Time  None of the Time | Single-select |
| 7 | How often during the past 2 weeks did you have trouble falling asleep? | All of the Time  Most of the Time  A Good Bit of the Time  Some of the Time  A Little of the Time  None of the Time | Single-select |
| 8 | How often during the past 2 weeks did you awaken during your sleep time and have trouble falling asleep again? | All of the Time  Most of the Time  A Good Bit of the Time  Some of the Time  A Little of the Time  None of the Time | Single-select |
| 9 | How often during the past 2 weeks did you have trouble staying awake during the day? | All of the Time  Most of the Time  A Good Bit of the Time  Some of the Time  A Little of the Time  None of the Time |  |
| 10 | How often during the past 2 weeks did you get the amount of sleep you needed? | All of the Time  Most of the Time  A Good Bit of the Time  Some of the Time  A Little of the Time  None of the Time | Single-select |

#### **PCL-5**

| **PCL-5:**  *Approximate time to complete: 10 min* | | | |
| --- | --- | --- | --- |
| # | Question | Response Options | Notes |
| N/A | These questions will help us learn more about you and your PTSD symptoms. We need to ask these questions the same way a doctor would, so you may have seen these questions before at the VA.  It’s ok if some of these questions are hard for you to answer. There are no “right” or “wrong” answers because everyone’s experience is unique to them. If you feel stuck or you are not sure how to answer a question, just do the best you can.  **Remember: How you respond to these questions will not go on your medical record or be reported to the VA, so please answer honestly. This information will be used only for this research study. Please see your Informed Consent Form for more information on how we keep your information private.** | N/A | Introduction |
| N/A | Instructions: Below is a list of problems that people sometimes have in response to a very stressful experience. Please read each problem carefully and then select one of the options to the right to indicate how much you have been bothered by that problem in the past month. | N/A | Introduction |
| 1 | In the past month, how much were you bothered by:  Repeated, disturbing, and unwanted memories of the  stressful experience? | Not at all  A little bit  Moderately  Quite a bit  Extremely | Single-select |
| 2 | In the past month, how much were you bothered by:  Repeated, disturbing dreams of the stressful experience? | Not at all  A little bit  Moderately  Quite a bit  Extremely | Single-select |
| 3 | In the past month, how much were you bothered by:  Suddenly feeling or acting as if the stressful experience were actually happening again (as if you were actually back there reliving it)? | Not at all  A little bit  Moderately  Quite a bit  Extremely | Single-select |
| 4 | In the past month, how much were you bothered by:  Feeling very upset when something reminded you of the  stressful experience? | Not at all  A little bit  Moderately  Quite a bit  Extremely | Single-select |
| 5 | In the past month, how much were you bothered by:  Having strong physical reactions when something reminded  you of the stressful experience (for example, heart  pounding, trouble breathing, sweating)? | Not at all  A little bit  Moderately  Quite a bit  Extremely | Single-select |
| 6 | In the past month, how much were you bothered by:  Avoiding memories, thoughts, or feelings related to the  stressful experience? | Not at all  A little bit  Moderately  Quite a bit  Extremely | Single-select |
| 7 | In the past month, how much were you bothered by:  Avoiding external reminders of the stressful experience (for  example, people, places, conversations, activities, objects, or  situations)? | Not at all  A little bit  Moderately  Quite a bit  Extremely | Single-select |
| 8 | In the past month, how much were you bothered by:  Trouble remembering important parts of the stressful  experience? | Not at all  A little bit  Moderately  Quite a bit  Extremely | Single-select |
| 9 | In the past month, how much were you bothered by:  Having strong negative beliefs about yourself, other people,  or the world (for example, having thoughts such as: I am  bad, there is something seriously wrong with me,  no one can be trusted, the world is completely dangerous)? | Not at all  A little bit  Moderately  Quite a bit  Extremely | Single-select  **A response of “Extremely” should trigger an alert to the on-call clinician** |
| 10 | In the past month, how much were you bothered by:  Blaming yourself or someone else for the stressful  experience or what happened after it? | Not at all  A little bit  Moderately  Quite a bit  Extremely | Single-select |
| 11 | In the past month, how much were you bothered by:  Having strong negative feelings such as fear, horror, anger, guilt, or shame? | Not at all  A little bit  Moderately  Quite a bit  Extremely | Single-select |
| 12 | In the past month, how much were you bothered by:  Loss of interest in activities that you used to enjoy? | Not at all  A little bit  Moderately  Quite a bit  Extremely | Single-select |
| 13 | In the past month, how much were you bothered by:  Feeling distant or cut off from other people? | Not at all  A little bit  Moderately  Quite a bit  Extremely | Single-select |
| 14 | In the past month, how much were you bothered by:  Trouble experiencing positive feelings (for example, being  unable to feel happiness or have loving feelings for people  close to you)? | Not at all  A little bit  Moderately  Quite a bit  Extremely | Single-select |
| 15 | In the past month, how much were you bothered by:  Irritable behavior, angry outbursts, or acting aggressively? | Not at all  A little bit  Moderately  Quite a bit  Extremely | Single-select |
| 16 | In the past month, how much were you bothered by:  Taking too many risks or doing things that could cause you  harm? | Not at all  A little bit  Moderately  Quite a bit  Extremely | Single-select  **A response of “Extremely” should trigger an alert to the on-call clinician** |
| 17 | In the past month, how much were you bothered by:  Being “superalert” or watchful or on guard? | Not at all  A little bit  Moderately  Quite a bit  Extremely | Single-select |
| 18 | In the past month, how much were you bothered by:  Feeling jumpy or easily startled? | Not at all  A little bit  Moderately  Quite a bit  Extremely | Single-select |
| 19 | In the past month, how much were you bothered by:  Having difficulty concentrating? | Not at all  A little bit  Moderately  Quite a bit  Extremely | Single-select |
| 20 | In the past month, how much were you bothered by:  Trouble falling or staying asleep? | Not at all  A little bit  Moderately  Quite a bit  Extremely | Single-select |

#### **Change in Sleep Management Questionnaire**

| **Change in Sleep Management:**  *Approximate time to complete: 5 min* | | | |
| --- | --- | --- | --- |
| # | Question | Response Options | Notes |
| N/A | These questions will help us understand if you’ve changed any regular habits in the last two weeks. Please answer No or Yes.  There are no “right” or “wrong” answers because everyone’s experience is unique to them. If you feel stuck or you are not sure how to answer a question, just do the best you can.  **Remember: How you respond to these questions will not go on your medical record or be reported to the VA, so please answer honestly. This information will be used only for this research study. Please see your Informed Consent Form for more information on how we keep your information private.** | N/A | Introduction |
| 1 | Do you use tobacco a lot more or less than you did before you started the Veterans ECS21 study? | No  Yes | Single-Select |
| 2 | *During run-in phase:*  Do you use cannabis a lot more or less than you did before you started the Veterans ECS21 study?  *During treatment phase:*  *Except* for using the study product, do you use cannabis a lot more or less than you did before you started the Veterans ECS21 study? | No  Yes | Single-Select |
| 3 | Have you changed any prescribed or over-the-counter medications or supplements you use (for example, stopping a medication, starting a new medication, or changing how much you take)? | No  Yes | Single-Select |
| 4 | Have you started or stopped any other sources of support to help you sleep (for example, sleep-focused therapy, meditation, exercise, other)? | No  Yes | Single-Select |
| 5 | Have you had any significant changes to your sleeping environment, such as:   - Moving to a new house - Rearranging furniture in your bedroom - Having a new baby, new pet, new construction noise nearby, etc that is interrupting your sleep? - Something you’ve changed about what’s normally on or off during sleep (such as a TV, radio, fan, noise machine, etc)? | No  Yes | Single-Select |

| **Daily Diary (Evening): Presented daily through the Run-In phase of the study**  *Approximate time to complete: 1 min* | | | |
| --- | --- | --- | --- |
| # | Question | Response Options | Notes |
| 1 | How satisfied are you with your day? | Very 😀  Somewhat 😐  A little bit 🙁  Not at all 😩 | Single-Select |
| 2 | Did you feel productive today? | No  Yes | Single-Select |

#### **Daily Diary**

#### **Daily Diary & Treatment Adherence Diary**

| **Daily Diary & ProTab^TM^ Adherence Diary (Evening): Presented daily through the Treatment phase of the study.**  *Approximate time to complete: 2 min* | | | |
| --- | --- | --- | --- |
| # | Question | Response Options | Notes |
| 1 | How satisfied are you with your day? | Very 😀  Somewhat 😐  A little bit 🙁  Not at all 😩 | Single-Select |
| 2 | Did you feel productive today? | No  Yes | Single-Select |
| 3 | Did you take your study product (ProTab^TM^) today? | No  Yes | Single-Select |
| 3a | Around what time did you take your study product (ProTab^TM^) today?  Just get as close to the right hour of the day as you can. | HH:MM AM/PM | **Only presented if they select “Yes” to Question 3** |
| 3b | How many tablets of the study product (ProTab^TM^) did you take? | 1  2  Other (free text) | **Only presented if they select “Yes” to Question 3** |

#### **WHODAS-2.0-12**

| **WHODAS-2.0-12**  *Approximate time to complete: 8 min* | | | |
| --- | --- | --- | --- |
| # | Question | Response Options | Notes |
| N/A | These questions will help us learn more about how you feel overall.  We need to ask these questions the same way a doctor would, so you may have seen these questions before at the VA.  It’s ok if some of these questions are hard for you to answer. There are no “right” or “wrong” answers because everyone’s experience is unique to them. If you feel stuck or you are not sure how to answer a question, just do the best you can.  **Remember: How you respond to these questions will not go on your medical record or be reported to the VA, so please answer honestly. This information will be used only for this research study. Please see your Informed Consent Form for more information on how we keep your information private.** | N/A | Introduction |
| N/A | This questionnaire asks about difficulties due to health conditions. Health conditions include diseases or illnesses, other health problems that may be short or long lasting, injuries, mental or emotional problems, and problems with alcohol or drugs.  Think back over the past 30 days and answer these questions, thinking about how much difficulty you had doing the following activities. | N/A | Introduction |
| 1 | In the past 30 days, how much difficulty did you have in:  Standing for long periods such as 30 minutes? | None  Mild  Moderate  Severe  Extreme or cannot do | Single-select |
| 2 | In the past 30 days, how much difficulty did you have in:  Taking care of your household responsibilities? | None  Mild  Moderate  Severe  Extreme or cannot do | Single-select |
| 3 | In the past 30 days, how much difficulty did you have in:  Learning a new task, for example, learning how to get to a new place? | None  Mild  Moderate  Severe  Extreme or cannot do | Single-select |
| 4 | In the past 30 days, how much difficulty did you have in:  How much of a problem did you have joining in community activities (for example, festivities, religious or other activities) in the same way as anyone else can? | None  Mild  Moderate  Severe  Extreme or cannot do | Single-select |
| 5 | In the past 30 days, how much difficulty did you have in:  How much have you been emotionally affected by your health problems? | None  Mild  Moderate  Severe  Extreme or cannot do | Single-select |
| 6 | In the past 30 days, how much difficulty did you have in:  Concentrating on doing something for ten minutes? | None  Mild  Moderate  Severe  Extreme or cannot do | Single-select |
| 7 | In the past 30 days, how much difficulty did you have in:  Walking a long distance such as more than half a mile? | None  Mild  Moderate  Severe  Extreme or cannot do | Single-select |
| 8 | In the past 30 days, how much difficulty did you have in:  Washing your whole body? | None  Mild  Moderate  Severe  Extreme or cannot do | Single-select |
| 9 | In the past 30 days, how much difficulty did you have in:  Getting dressed? | None  Mild  Moderate  Severe  Extreme or cannot do | Single-select |
| 10 | In the past 30 days, how much difficulty did you have in:  Dealing with people you do not know? | None  Mild  Moderate  Severe  Extreme or cannot do | Single-select |
| 11 | In the past 30 days, how much difficulty did you have in:  Maintaining a friendship? | None  Mild  Moderate  Severe  Extreme or cannot do | Single-select |
| 12 | In the past 30 days, how much difficulty did you have in:  Your day-to-day work? | None  Mild  Moderate  Severe  Extreme or cannot do | Single-select |

#### **Sleep Diary**

| **Sleep Diary (Morning):**  *Approximate time to complete: 4 min* | | | |
| --- | --- | --- | --- |
| # | Question | Response Options | Notes |
| 1 | How was your sleep last night? | Very good 😴  Good 🙂  Okay 😐  Poor 🙁  Very Poor 🥱 | Single-select |
| 2 | Did you set an alarm to wake up this morning? | No  Yes | Single-select |
| 3 | Are you going to work today? | No  Yes | Single-select |
|  | These questions will help us learn about how your activities last night might have affected your sleep.  **Remember: How you respond to these questions will not go on your medical record or be reported to the VA, so please answer honestly. This information will be used only for this research study. Please see your Informed Consent Form for more information on how we keep your information private.** | N/A | Introduction |
| 4 | Last night, did you have a nightmare that interrupted your sleep? | No  Yes | Single-select |
| 5 | Did any other unusual events affect your sleep last night (for example, an emergency, sickness, travel)? | No  Yes | Single-select |
| 6 | Last night, did you have a drink containing alcohol? | No  Yes  I’m not sure | Single-select |
| 6a | How many drinks containing alcohol did you have last night? | 1 or 2  3 or 4  5 or 6  More than 6  I’m not sure | **Only presented if they select “Yes” to Question 6**  Single-select |
| 7 | Last night, did you use drugs?  The various classes of drugs may include: cannabis (e.g., marijuana, hash), solvents, tranquillizers (e.g., Valium), barbiturates, cocaine, stimulants (e.g., speed), hallucinogens (e.g., LSD) or narcotics/opioids (e.g., heroin, fentanyl, oxycodone - oxyz). | No  Yes  I’m not sure | Single-select |
| 8 | Last night, did you take a sleep aid (a prescribed or over-the-counter medication or supplements for sleep?)  For example: Ambien, Zolpidem, Lunesta, benzodiazepines, unisom, diphenhydramine, doxylamine, melatonin, valerian, etc? | No  Yes  I’m not sure | Single-select |
| 9 | Did you do anything last night to specifically help you relax right before bed (like meditation, yoga, reading, therapy, etc)? | No  Yes | Single-select |

#### **ProTab^TM^ Feedback**

| **ProTab^TM^ Feedback:**  *Approximate time to complete: 3 min* | | | |
| --- | --- | --- | --- |
| # | Question | Response Options | Notes |
| N/A | These questions will help us understand your experience with the study product (ProTab^TM^). | N/A | Introduction |
| 1 | It was easy for me to swallow the ProTab™ | Strongly disagree  Disagree  Neutral  Agree  Strongly agree | Single-select |
| 2 | How soon after taking the Protab™ did you feel an effect? | Within 15 minutes  Within an hour  Within a few hours  I’m not sure  I didn’t feel an effect | Single-select |
| 3 | Was the effect of the Protab™ pleasant or unpleasant? | Very unpleasant  Unpleasant  Pleasant  Very pleasant  I’m not sure  I didn’t feel an effect | Single-select |

#### **Study Feedback**

| **Study Feedback:**  *Approximate time to complete: 5 min* | | | |
| --- | --- | --- | --- |
| # | Question | Response Options | Notes |
| N/A | Thanks for being part of our study! These questions will ask about your overall study experience and collect final study data. | N/A | Introduction |
| 1 | What is your current weight (in pounds)? | NNN | Integer |
| 2 | Overall, I am satisfied with my participation in the study. | Strongly disagree  Disagree  Neutral  Agree  Strongly agree | Single-select |
| 3 | Learning how to use the study application was easy. | Strongly disagree  Disagree  Neutral  Agree  Strongly agree | Single-select |
| 4 | I plan to use the Fitbit after the study is over. | Strongly disagree  Disagree  Neutral  Agree  Strongly agree | Single-select |
| 5 | It was easy for me to find the time to do the study activities. | Strongly disagree  Disagree  Neutral  Agree  Strongly agree | Single-select |
| 6 | On days you did not complete the study activities (questionnaires or calls with the research team), what was your most common reason for not doing them? | I didn’t have time  There were too many to do  The surveys made me uncomfortable  It was difficult to answer the surveys in the app  Other  Does not apply to me - I completed all the surveys | Single-select |
| 7 | On a scale of 0 (very unlikely) to 10 (very likely), would you recommend participating in this study to a friend? | 0-10 | Single-select |
| 8 | Do you plan to continue taking CBG after the study is over? | Yes  No  I’m not sure | Single-select |
| 9 | What can we do to improve the study experience for future participants? | *N/A* | Free text |
| 10 | What was your favorite feature of the study? | *N/A* | Free text |

### **Supplementary Table 4: CRC Guide for Check-In Calls**

**Recommendations for Tricky Topics**

**Establishing rapport:** Open the conversation with a casual and personalized prompt. You can ask how their week has been, how they have been feeling since you last talked, how work is going (if they work), or ask about something they brought up in a prior conversation like their kid(s) or pet(s), if those topics feel casual and friendly enough.

**Compliance Issues:** We have noticed that we are not getting as much data from the surveys as we expected. We are here to support you and we want to help you however we can. Where are you getting stuck? *[Listen & document study feedback, clarify any questions they have]*

*ONLY if it seems like they are really disengaged and unwilling to participate*: Do you want to continue your participation in the study?

*[If no]:* If it is just not fitting into your life at the moment that is completely understandable, and we are so appreciative of the fact that you wanted to be a part of this research. We’d love to get some feedback to help us understand how to make the study better. If you could change something about the study, what would you change that you think would have made it easier for you?

**Privacy:** Any information you share with me or in the study application is just for the research team. It won’t be reported to the VA or go on your medical record.

**Honesty:** There are no right or wrong answers -- everyone is different, and we’re doing this study to help Veterans like you. Any information you share with me or in the study application is just for the research team. It won’t be reported to the VA or go on your medical record, so we want to encourage you to answer honestly.

**Getting “stuck”:** I know some of the questions can be hard to answer. Just remember that if you start to feel stuck, just do the best you can.

| **Consent Call** | |
| --- | --- |
|  | **Introduce the study & your role**  Suggested language: *“We want to see if CBG (a type of hemp) can help Veterans sleep better. This study is sponsored by a Veteran-founded company and we want to find new ways to help Veterans. Thank you for your service and your interest in the study.”* |
|  | **Summarize study structure**  Suggested language: *“This is a 6-week study. During the first two weeks, we’re just gathering information about your normal sleep. Then you’ll start taking 1 tablet of the study product daily for two weeks, and then you’ll take 2 tablets of the study product daily for two weeks. All the materials you need will be shipped directly to you.”* |
|  | **Summarize study activities**   - Answer questionnaires (twice daily, plus 1-2 additional questionnaires each week). This should take you about 5 minutes a day, on average. - Wear a Fitbit during the day and overnight. - Have a call with [me/the study team] every other week |
|  | **Medication expectations**  Suggested language: *“We won’t ask you to change any medications you’re currently taking, but we do want you to tell us if you stop, start, or change dosages because that might impact how you’re sleeping.”* |
|  | **Consent:** Go through consent & answer any additional questions |
|  | **Collect contact information** |
|  | **Schedule next call:** This call should occur 4-8 days from today |

| **Fitbit & App Onboarding Call (Day 0)**  Goals:   - Establish rapport - Ensure participant is fully set up with the study application & Fitbit | |
| --- | --- |
|  | Emphasize privacy & your role for support |
|  | **App Onboarding:** Go through App section of the Participant Guide. Confirm they see 2 questionnaires (Participant Info & PCL-5). |
|  | **Remind About Survey Windows:** Remind participant they have until end of day tomorrow to complete the questionnaires. It should take about 10 minutes.  Suggested language: *“I know some of the questions can be hard to answer. Just remember that if you start to feel stuck, just do the best you can.”* |
|  | **Ongoing App Surveys**  Suggested language: *“Starting tomorrow, you’ll see two questionnaires to complete each day (one in the morning, one in the evening). These should only take a few minutes to complete. You’ll see some additional questionnaires once or twice a week that take a little more time (5-10 minutes). We estimate you’ll spend about 5 minutes a day.”* |
|  | **Prompt habit-setting behavior:** Examples: Setting an alert on their phone, post-it note on the mirror, etc...  Suggested language: *“We decided not to have the app remind you every day to answer your questionnaires. What is a good way for you to remember to do this each day?* |
|  | **Fitbit Onboarding:** Go through Fitbit section of the Participant Guide |
|  | **Fitbit Expectations:**  Suggested language: *“Starting today, start wearing your Fitbit during the day and overnight. We recommend charging it while you shower.”* |
|  | **Schedule next check-in call**: Can occur Day 10 - Day 14 |

| **Check In Call (Day 14)**  Goals:   - Establish rapport & continue relationship development - Learn if participant has changed anything that might impact their sleep - Ensure participant is fully set up with the study product & what to expect | |
| --- | --- |
|  | **Friendly Opener** |
|  | **Study Feedback/Technical Issues**  *Suggested questions:*   - What have you thought about your study experience so far? - What do you think about the study application and surveys? - Have you had any issues with the study application or the Fitbit? |
|  | **Compliance** |
|  | **Changes in Sleep Management:**  Suggested language: *“I’d like to hear more about what’s changed and how it might have affected your sleep.”*  Gather more information about what has changed, including:   - What changed - How did it change - When did it change - Did they notice any changes to their sleep after the change? |
|  | **Study Product Onboarding**  Suggested language: *“Next week on [day], the [date], you’ll start taking the study product. As a reminder, you were randomly assigned by chance (like the flip of a coin) to receive either CBG or a placebo that doesn’t have any CBG. They’ll look the same, so you won’t know which one you’ve gotten.*  *When you start taking the study product on [day], the [date], swallow one tablet anytime after when you wake up and no later than 3 hours before you go to bed. You can take it with or without food.*  *Would it be helpful to set a reminder on your calendar on [day], [month date] or write the date on the card you got in your box?*  *One thing that could also help is to put the card near something that’s already a regular habit, like by your toothbrush or on your kitchen counter. What do you think would help you remember to take the study product each day?* |
|  | **New questions in app**  Suggested language: *“The day you start taking study product, you’ll see some new questions in the application. This is how you can let us know if you did or didn’t take your study product & what time you took it.”* |
|  | **Emphasize MOS Sleep Problems Index II:** This questionnaire is an important data point for the study. The participant needs to complete it on Day 14 or Day 15. |
|  | **Schedule next check-in call**: Can occur Day 24 - Day 28 |

| **Check In Call (Day 28)**  Goals:   - Maintain rapport - Learn if participant has changed anything that might impact their sleep - Ensure participant is prepared to change dosing | |
| --- | --- |
|  | **Friendly Opener** |
|  | **Study Feedback/Technical Issues:**  *Suggested Questions:*   - What do you like the most about being in the study? - What do you like the least or what do you wish was different about the study? - Have you had any issues with the study application or the Fitbit? |
|  | **Compliance** |
|  | **Changes in Sleep Management:** Gather more information about what has changed, like:   - What changed - How did it change - When did it change - Did they notice any changes to their sleep after the change? |
|  | **Study Product: Changing to 2 tablets daily dosing**  *Suggested language:*  Next week on *[day]*, *the [date]*, you’ll start taking 2 tablets of the study product instead of one tablet each day. Take the study product anytime after when you wake up and no later than 3 hours before you go to bed. You can take it with or without food.  Would it be helpful to set a reminder on your calendar on *[day]*, *the [date]* or write the date on the card you got in your box to help you remember when to change your dose? |
|  | **Emphasize MOS Sleep Problems Index II:** This questionnaire is an important data point for the study. The participant needs to complete it on Day 28 or Day 29. |
|  | **Schedule next check-in call**: Can occur Day 38 - Day 42 |

| **Check In Call (Day 42): Exit Interview**  Goals:   - Learn if participant has changed anything that might impact their sleep - Gather participant’s feedback on study & ensure they feel appreciated | |
| --- | --- |
|  | **Friendly Opener** |
|  | **Compliance** |
|  | **Changes in Sleep Management:** Gather more information about what has changed, like:   - What changed - How did it change - When did it change - Did they notice any changes to their sleep after the change? |
|  | **Study Feedback/Technical Issues:**   - What do you like the most about being in the study? - What do you like the least or what do you wish was different about the study? - Have you had any issues with the study application or the Fitbit? - If you could change something about the study, what would you change that would have made it easier for you? - Do you think you’d participate in another research study? Why/why not? - Is there anything else you’d like us to know? |
|  | **Emphasize MOS Sleep Problems Index II, WHODAS, and PCL-5:** These questionnaires are the most important data points for the study.   - MOS Sleep Problems Index II: Day 42 or 43 - WHODAS & PCL-5: Day 43 or 44 |
|  | **End on a positive!**  Suggested language: *“We will use this information to better understand how CBG changes the way you sleep. This research will help us find new ways to help Veterans. You have helped us so much by participating in the study! Thank you for sharing your experiences & thank you for your service.”* |

### **References for Supplementary Material**

1.	Cannabis Data Company (BDSA). Retail sales tracking. Share of Market cannabinoids by sales (Jan, 2021 through June, 2022). Available at: <https://bdsa.com/products/retail-sales-tracking/>. [Accessed 8 July 2022].
